# A novel multi-omics-based identification of symptoms, comorbid conditions, and possible long-term complications in COVID-19

**DOI:** 10.1101/2020.12.08.20245753

**Authors:** Debmalya Barh, Sandeep Tiwari, Bruno Silva Andrade, Marianna E. Weener, Aristóteles Góes-Neto, Vasco Azevedo, Preetam Ghosh, Nirmal Kumar Ganguly

## Abstract

Till date the comprehensive clinical pictures, comorbid conditions, and long-term complications of COVID-19 are not known. Recently using a multi-omics-based strategy, we have predicted the drugs for COVID-19 management with ∼70% accuracy. Here, using a similar multi-omics-based bioinformatics approach and three-ways of analysis, we identified the symptoms, comorbid conditions, and short, mid and possible long-term complications of COVID-19 with ∼90% precision. In our analysis (i) we identified 27 parent, 170 child, and 403 specific conditions associated with COVID-19. (ii) Among the specific conditions, 36 are viral and 53 short-term, 62 short to mid to long-term, 194 mid to long-term, and 57 are congenital conditions. (iii) At a cut off “count of occurrence” of 4, we found ∼ 90% of the enriched conditions are associated with COVID-19. (iv) Except the dry cough and loss of taste, all other COVID-19 associated mild and severe symptoms are enriched. (v) Cardiovascular, pulmonary, metabolic, musculoskeletal, neuropsychiatric, kidney, liver, and immune system disorders are found as top comorbid conditions. (vi) Specific diseases such as myocardial infarction, hypertension, COPD, lung injury, diabetes, cirrhosis, mood disorders, dementia, macular degeneration, chronic kidney disease, lupus, arthritis etc. along with several other diseases are also enriched as top candidates. (vii) Interestingly, many cancers and congenital disorders associated with COVID-19 severity are also identified. (viii) Arthritis, dermatomyositis, glioma, diabetes, psychiatric disorder, cardiovascular diseases having bidirectional relationship with COVID-19 are also found as top ranked conditions. Based on the accuracy (∼90%) of this analysis, long presence of SARS-CoV-2 RNA in human, and our previously proposed “genetic remittance” assumption, we hypothesize that all the identified comorbid conditions including the short-long-mid and mid-long non-communicable diseases (NCDs) could also be long-term consequences in COVID-19 survivors and warrants long-term observational studies.

## INTRODUCTION

Since the first report of SARS-CoV-2 infection between December 7, 2019 to December 12, 2019 from China [1-3], as of November 10, 2020, the virus has infected 49,727,316 people with 1,248,373 deaths globally [4].

### Symptoms and comorbid conditions

Recent systematic reviews and meta-analysis based studies show that the **most common short-term clinical symptoms** in COVID-19 include fever (81.2%-84.30%), cough (58.5%-63.01%), fatigue (34.22% - 38.5%), dyspnea (26.1% −37.16%), Sputum (25.8%), diarrhea (11.47%) [5, 6], myalgia (7.7%), nasal congestion (17%), abdominal pain (2.4%), and emotional disturbances (15.9%) [7]. However, in critical cases, the most common complications are acute respiratory distress syndrome (ARDS) (33.15%), arrhythmia (16.64%), acute cardiac injury (15.68%), heart failure (11.50%), and acute kidney injury (AKI) (9.87%) [6]. Mortality and severity of COVID-19 is associated with various factors including gender (male, OR:1.60), behaviour (current smoker, OR:2.06), and existing several comorbid conditions [8].

**The most important comorbid conditions** are hypertension (OR:2.05), diabetes (OR:2.46), coronary heart diseases (OR:4.10), chronic kidney diseases (OR:4.06), and cancers (OR:2.28) [8]. A meta-analysis on underlying comorbid conditions in expired COVID-19 patients reveals hypertension is the most common comorbidity (46%) in COVID-19 followed by diabetes (26%), cardiovascular diseases (21%), cerebrovascular disease (13%), cancers (11%), Lung disease (11%), COPD (8%), kidney disease (7%), liver disease (3%), and asthma (3%) [9]. However, another meta-analysis suggests that asthma as a comorbid condition may not increase the mortality (OR: 0.96) [10]. Increased aspartate aminotransferase is associated with high severity and death from COVID-19 (OR: 4.48) [11] and patients with cirrhosis showed worsening liver function and increased mortality [12]. Adverse drug-drug interactions may also potentially worsen the outcomes in comorbid conditions such as cardiometabolic diseases, chronic kidney disease, and COPD [13]. Other identified increased risk factors are polycystic ovarian disease, vitamin D deficiency, and obesity [14] and the list is long.

Clark et al. based on a Global Burden of Diseases, Injuries, and Risk Factors Study (GBD) 2017 has provided a list of comorbid conditions that may increase the risk of COVID-19 severity and fatality. The conditions include HIV/AIDS, tuberculosis, various cancers cardiovascular diseases, chronic respiratory diseases, chronic liver diseases, diabetes, chronic kidney diseases, chronic neurological disorders, and sickle cell disorders [15].

Mudatsir and colleagues in their systematic review and meta-analysis analysed 62 potential risk factors and have suggested that, severe COVID-19 is associated with more than 30 risk factors. These factors include comorbid conditions such as hypertension, diabetes, and chronic respiratory and cardiovascular diseases. In respect to symptoms, fatigue, dyspnea, increased respiratory rate, anorexia, and high systolic blood pressure are associated with severity of COVID-19. Apart from these, a higher level of blood troponin, aspartate aminotransferase, alanine aminotransferase, lactate dehydrogenase, procalcitonin, C-reactive protein, D-dimer, creatinine, ferritin, urea, interleukin, ESR and low levels of lymphocytes and haemoglobin also correlate with the disease severity from COVID-19 [16]. Further, decreased lymphocyte and platelet counts, and elevated D-dimer fibrin degradation are associated with poor prognosis [17].

### Long- haul COVID-19

Apart from the disease severity, recently long-haul COVID-19 is also reported frequently. Although most of the COVID-19 symptoms disappear within 21 days, nearly 10% of COVID-19 patients show symptoms even after three weeks, 5% for eight weeks or more, and 2% suffer for nearly three months [18].

As discussed, several extra-pulmonary manifestations are found in COVID-19 [19]. Report suggests that the SARS-CoV-2 viral RNA remain present at low copies in various body samples of patients even after 75 days from the start of symptoms, suggesting the ability of this virus to provoke persistent infection and induce extra-pulmonary clinical symptoms [20]. It is reported that, only 10.8% COVID-19 patients do not show any post-COVID manifestation after recovery from the disease [21]. However, several symptoms persist even after 3 months. The most common post-COVID symptoms are fatigue and dyspnea. Other frequent complications include joint and chest pain, myocardial inflammation, myocarditis, pulmonary dysfunction, pulmonary fibrosis, headache, vertigo, anosmia, ageusia, encephalitis, seizures, mood disorder and “brain fog”,[22] lack of concentration, confusion, memory loss, dizziness, heart palpitations, tachycardia, and pain with deep breaths [23]. Low lung diffusion capacity is observed in 42% cases, besides also low exercise capacity (22% cases), mental and/or cognitive function (36% cases), fatigue (69%-72.8% cases) [21, 24], and headache (37.8% cases) [25] among others. Other less frequent post-COVID symptoms so far reported are acalculous cholecystitis [26], dry eye disease [27], sudden onset sensorineural hearing loss [28], stroke, renal failure [21], autoimmune orthostatic hypoperfusion syndrome/painful small fiber neuropathy [29] etc. It is also found that all COVID-19 survivors are at risk of developing the long-term complications [23]. Current evidence suggests that COVID-19 patients are at higher risk in developing mental health problems that include depression, anxiety disorders, stress, panic attack, irrational anger, impulsivity, somatization disorder, sleep disorders, emotional disturbance, posttraumatic stress symptoms, and suicidal behaviour [30].

### Long-term consequences of COVID-19

According to WHO, COVID-19 may increase the risk of developing long-term health issues related to cardiovascular systems, pulmonary system, brain and nervous system, musculoskeletal system, and mental health [31]. Apart from these, damage to kidney, liver and other endocrine system organs were also recently found in COVID-19 survivors [32]. Further, bidirectional relationship between COVID-19 and diabetes [33], psychiatric [34], and cardiovascular diseases [35] are also reported. Since the disease is just a year old, it is not yet well understood what could be the most common long-term health complications in COVID-19 survivors.

Based on this background, we hypothesize that, presence of comorbid conditions is associated with increased severity and fatality in COVID-19 and COVID-19 survivors may develop these comorbid conditions as long-term complications of COVID-19. So far, epidemiology, meta-analysis, case series, large cohort studies, review, GBD-2017, and longitudinal dynamics based approach have been used to identify the comorbid conditions and long-term complications in COVID-19. However, omics-based approach has not been used for prediction or identification of these conditions.

Recently, applying a multi-omics based bioinformatics analysis; we have predicted the SARS-CoV-2 infection biology, the deregulated pathways, and potential drugs for COVID management with 70% precision [36]. Assuming that the diseases/ signs / symptoms associated with these upregulated multi-omics datasets are (i) related with COVID-19, (ii) contribute to COVID-19 associated comorbid conditions or risk factors, (iii) correlate with general or severe signs and symptoms of COVID-19, and (iv) these comorbid conditions could also be the long-term complications of COVID-19; in this analysis, we have used the same datasets (interactome, proteome, transcriptome, and bibliome) used in our previous report and a modified multi-omics approach [36] to predict the comorbid conditions, signs and symptoms, and the long-term complications in COVID-19.

## METHODS

### Datasets

Similar to our previous work [36], we have used the same five omics data sets: interactome data (I) of Gordon et al., 2020 [37], proteome data (P) of Bojkova et al., 2020 [38], transcriptome profiles (T-1 and T-2) of Blanco-Melo et al., 2020 [39] and Xiong et al., 2020 [40] along with the same bibliome data (B). Only the upregulated genes or proteins were considered for analysis from the datasets used in this work. Individual omics data (I, P, T-1, T-2, B) and their three combinations were used. The combinations were P+T (P+T-1+T-2), P+T+I, and P+T+I+B.

### Data analysis

#### Gene-set based disease enrichment

Like our previous analysis, we have used the ToppFun and ToppGene modules of the ToppGene suite (https://toppgene.cchmc.org) with default parameters [41] for disease enrichment. In case of ToppGene module, the bibliome dataset (B) of 745 COVID-19 related genes were used as training set. The second enrichment tool used was Enrichr (https://amp.pharm.mssm.edu/Enrichr/) [42]. Six different databases integrated with the Enrichr [42] were selected to enrich the diseases or conditions. The GEO up, PheWeb, DisGeNET databases along with GWAS Catalog, OMIM Expanded, and Rare Diseases were used. The GWAS Catalogue, OMIM Expanded, and Rare Diseases based analysis were performed keeping in mind the “genetic predisposition” hypothesis (discussed later) to predict the possible genetic diseases associated with the gene sets used. Based on the P-value, Odds ratio, and combined score, the top 20 diseases from each analysis were then combined for a consolidated analysis.

#### Consolidated disease analysis

Contrary to our previous analytic approach [36], we have applied some additional considerations in this analysis. We have considered one of our previous hypotheses, i.e. “genetic remittance” [43]. According to this hypothesis, a virus or bacteria leaves a permanent signature inside host upon infection and that the host carries the signature for a long time. Therefore, pathways that are activated during the infection remains switched on for a considerable duration within the host and subsequently develop clinical symptoms in long term. The RNA of the SARS-CoV-2 remains active in the COVID-19 survivors for more than 75 days [20] and thus the virus can maintain the upregulation of these omics profile for long periods and as long-term consequences of it, the non-communicable disease (NCD) pathways associated with these upregulated omics profile also remain active and subsequently will trigger the development of the clinical symptoms/phenotypes in long term.

In our gene-set-based disease enrichment, several unique terminologies for a disease or clinical symptom were enriched. Therefore, in the consolidated disease analysis, first we classified the enriched diseases or symptoms based on Disease Ontologies using Ontobee tool (http://www.ontobee.org) [44]. In this process, first we classified each disease or symptom under three relationships (parent - child - specific disease/symptom). For example, the “hypertensive disease” identified in gene-set-based disease enrichment is classified as cardiovascular system disease - vascular disease - hypertensive disease. Next, we grouped and counted each parent – child - specific disease/symptom based on parent, child, and specific disease/ symptom individually.

Further, based on the time or duration it takes to develop and present their clinical pictures in normal condition, each specific disease/ sign / symptom is further classified under short, short-mid, mid-long, short-mid-long, short-mid-long-genetic, mid-long-genetic, and congenital/genetic. For example, fever is classified under short and liver cirrhosis could be tagged with mid-long. Finally, based on the cumulative number of occurrence (the score), the diseases/conditions are ranked and the final results are considered based on this rank. A cut off 4 on the occurrence score was considered in most of the analysis to select the disease or symptoms. It should be noted that, a particular cumulative score for two or more different diseases are considered to have the same rank although they are listed one by one and diseases having same score have equal probability. The overall approach applied in our work is presented in **Fig-1**.

**Fig.1:**
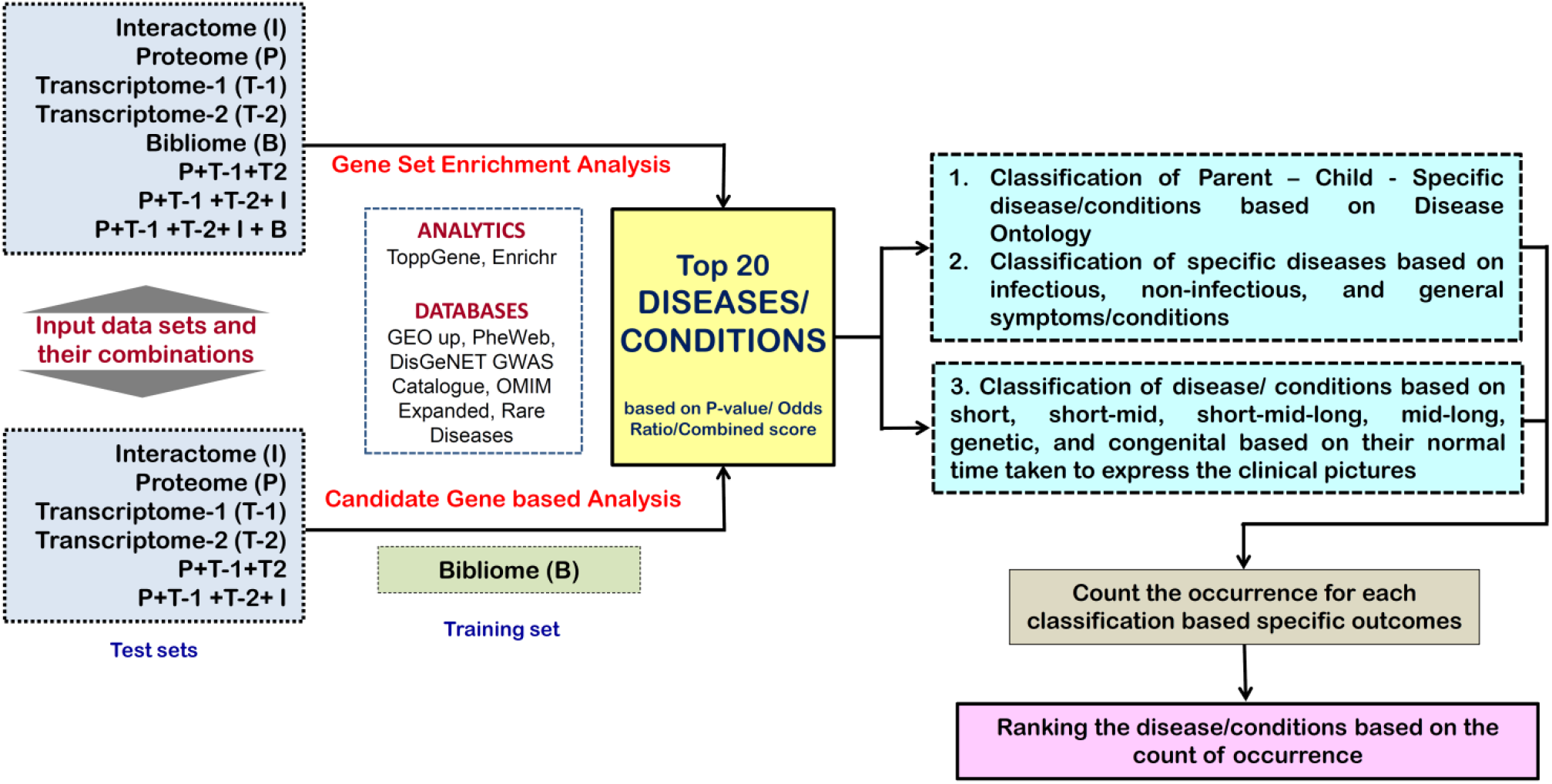
The flow diagram of the overall strategy applied in this analysis. For details, see the methods section.

## RESULTS AND DISCUSSION

We generated 1210 classified parent - child - specific disease/symptom associations based on our top 20 enriched diseases for various omics datasets and their combinations using TopGene, Enricher, and Ontobee [44]. While we cleaned the data removing the duplicate of a specific disease or symptom and merged the similar diseases or symptoms to make the counting calculations, we obtained a dataset having 403 specific diseases or symptoms and used this dataset for further analysis.

### COVID-19 associated parent disease or symptom groups

We identified 27 parent diseases or symptoms based on our consolidated analysis without classifying them based on the time duration needed to develop these diseases or symptoms. Infectious disease came at Rank-1, neoplasm (Rank-2), abnormal sign or symptom (Rank-3), cardiovascular system disease (Rank-4), immune system disorder (Rank-5), nervous system disease (Rank-6), musculoskeletal system disease (Rank-7), metabolic disorder (Rank-8), and mental disorder (Rank-9). At Rank-10 two diseases, namely, congenital anomalies and respiratory system disease are enriched. Eye disease is Ranked at 11, gastrointestinal system disease (Rank-12), urinary system disease (Rank-13), skin disease (Rank-14), liver disease (Rank-15), hematopoietic system disease (Rank-16), sepsis (Rank-17), Response to drug (Rank-18), behaviour process (Rank-19), and female reproductive system disease (Rank-19) constitute the top twenty disease classes (**Fig-2A**).

**Fig-2:**
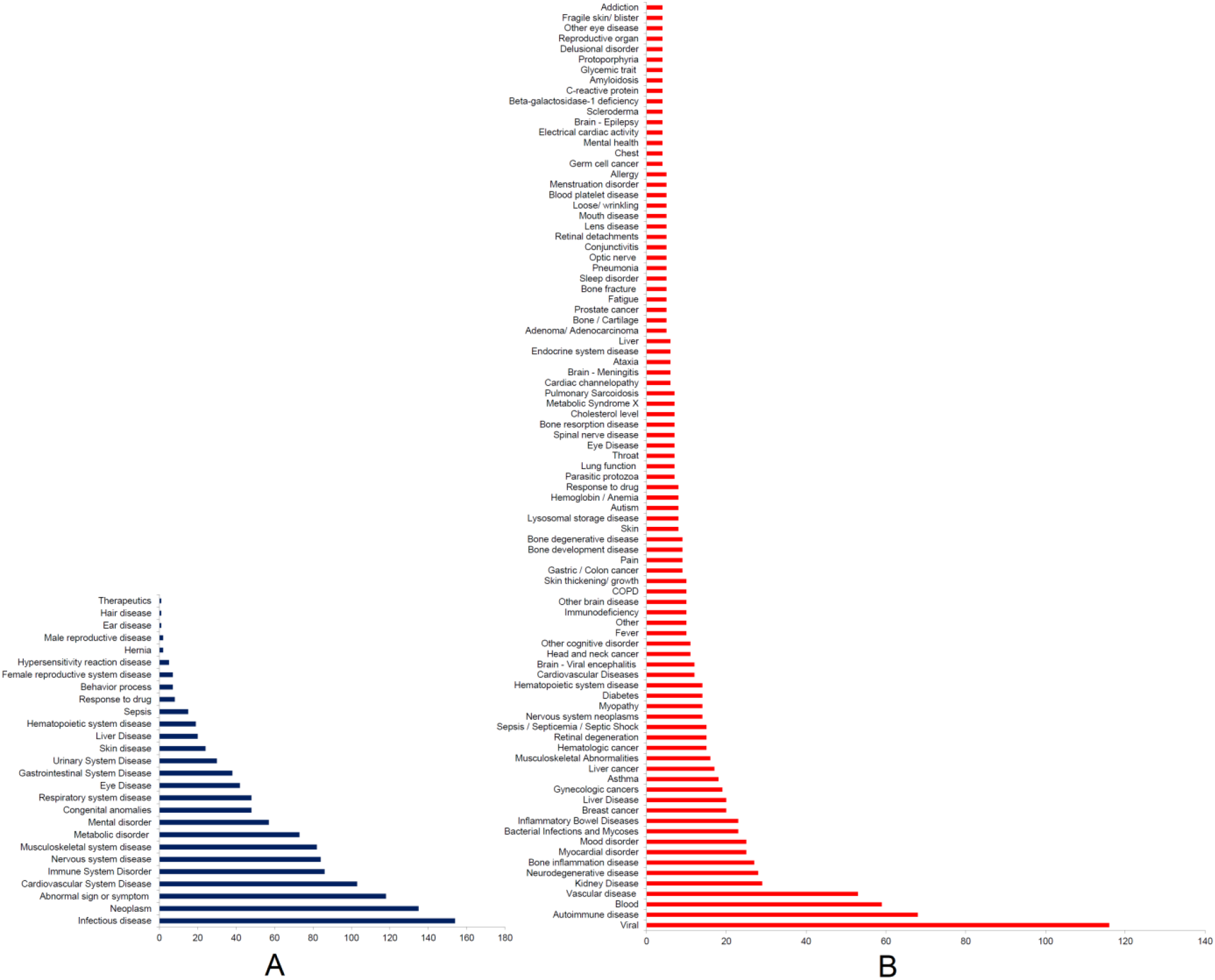
**A)** The top 27 parent diseases or symptoms as per the consolidated analysis. **B)** Top child diseases.

We found all most all the parent diseases as described by Clark et al. [15]. Our identified all the 27 parent diseases or symptoms are highly associated with COVID-19 which itself is a highly infectious disease [45]. Cancers [46], cardiovascular diseases [47, 48], autoimmune diseases [49, 50], metabolic disorders [51, 52], congenital heart disease [53, 54], respiratory system disease [48], gastrointestinal diseases [55], kidney diseases [56, 57], liver disease [58], hematopoietic system disease [59], sepsis or septicaemia [60] are the reported key comorbid conditions in COVID-19 that are associated with an increased susceptibility, disease severity and mortality from COVID-19.

Considerable neurological [61, 62], musculoskeletal [63, 64], mental or psychological or behaviour process [30, 34], reproductive system [65, 66] Eye diseases [67], and skin disease [68, 69] manifestations are observed during the infection and may have long-term complications. A higher preterm labour in pregnant women infected with COVID-19 [70] is also reported. However, we did not find any literature supporting major congenital anomalies due to COVID-19. Therefore, the prediction of the parent disease groups is nearly 90% accurate in our analysis.

### COVID-19 associated child diseases or conditions

While we analysed for the important child diseases or conditions under each parent disease, we observed that, viral diseases constitute 75% of the diseases followed by bacterial infections under the parent infectious diseases. For neoplasm, liver, breast, gynaecologic, neurologic, and head and neck cancers are highly enriched. The key abnormal sign or symptoms are associated with blood followed by fever, pain, lung function and throat infection, fatigue, mental health, chest, and eye diseases. Under cardiovascular system disease, vascular diseases are the most frequent followed by myocardial disorders. The autoimmune diseases followed by immunodeficiency are the key diseases under the parent immune system disorder. Neurodegenerative disease, viral encephalitis, and other cerebral diseases are the top three conditions under nervous system disease category. The top three conditions under musculoskeletal system disease are bone inflammation disease, myopathy, and bone degenerative disease. Diabetes, lysosomal storage disease, metabolic syndrome X, and cholesterol level are top candidates under metabolic disorder. For mental disorder, mood disorder is most frequent followed by cognitive disorder, autism, and sleep disorder. Under respiratory system disease, asthma, COPD, pulmonary sarcoidosis, pneumonia, and lung injury are the top 5 conditions. Inflammatory bowel diseases and kidney disease are respectively top conditions under gastrointestinal system and urinary system diseases, respectively. Therefore, this child disease analysis is also nearly 90% accurate if we compared with the symptoms and comorbid conditions of COVID-19 described in various literatures. The details of this analysis are given in **Fig-3**.

**Fig-3:**
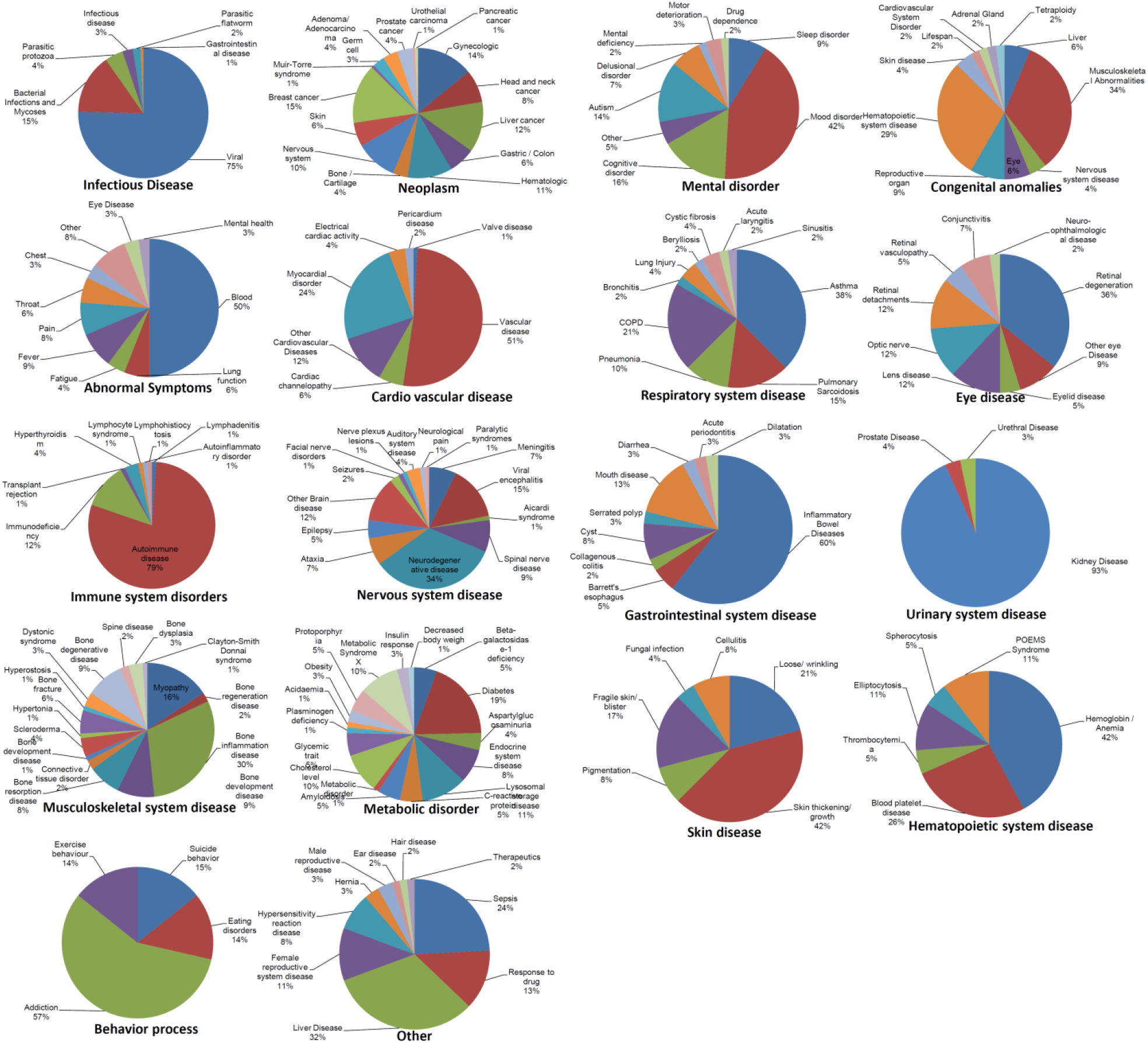
The proportion of child diseases under each parent disease

Apart from the above approach for child disease or symptoms analysis, we performed another enrichment analysis without considering the parent disease group (**Fig-2B**). In this analysis, we directly enriched the top child conditions based on their number of occurrences from the 404 classified parent - child - specific disease/symptom associations. In this analysis too, the viral infection is ranked one. The common comorbid conditions such as autoimmune disease (Rank-2) [50], vascular disease (Rank-4) [71], kidney disease (Rank-5) [56, 57], neurodegenerative disease (Rank-6) [72], bone inflammation disease (Rank-7) [73], myocardial disorder (Rank-7) [74], mood disorder (Rank-7) [34], breast cancer (Rank-10) [75], liver disease (Rank-10) [58], gynaecologic cancers (Rank-11) [76], asthma (Rank-12) [77], liver cancer (Rank-13) [78], musculoskeletal abnormality (Rank-14) [63], hematologic cancer (Rank-15) [79], retinal degeneration (Rank-15) [80], sepsis / septicaemia / septic shock (Rank-15) [60, 81], diabetes (Rank-16) [82], other cardiovascular diseases (Rank-17) [71, 74], viral encephalitis (Rank-17) [83], other cognitive disorder (Rank-18) [24], immunodeficiency (Rank-19) [84], brain disease (Rank-19) [85], COPD (Rank-19) [86], gastric / colon cancer (Rank-20) [87], autism (Rank-21) [88, 89], anemia (Rank-21) [90], bone development and degenerative disease (Rank-20) [91], dyslipidaemia (Rank-22) [92, 93], and ataxia (Rank-23) [94] etc. are among the top 25 child disease categories at a cut off score of 4 (**Fig-2B**). Inflammatory bowel disease which may not be a comorbid condition in COVID-19 [95] is found enriched (Rank-9). Importantly, nervous system neoplasms (Rank-16), myopathy (Rank-16), hematopoietic system disease (Rank-16), head and neck cancer (Rank-18), and pulmonary sarcoidosis (Rank-22) also came as top child diseases.

Apart from these diseases, general symptoms of COVID-19 such as, blood related abnormalities (Rank-3), fever / severe fever with thrombocytopenia syndrome (Rank-19) [5], pain (Rank-20) [18], lung/ pulmonary function (Rank-22) [96, 97], fatigue (Rank-24) [6], sleep disorder (Rank-24) [98], pneumonia [86], conjunctivitis (Rank-24) [99], skin diseases etc. are also enriched under top 25 child categories with occurrence cut off value of 4. We also found many other reported COVID-19 associated diseases/symptoms enriched below the cut off 4. For the details on the enriched child diseases/symptoms please see the **Fig-2B**.

Therefore, we achieved nearly 90% precision in identifying the diseases or symptoms associated with COVID-19 and hence the multi-omics based method we have developed and applied here is highly accurate and can be applied to other diseases too.

### COVID-19 associated specific diseases and symptoms

In this analysis, first we ranked the specific diseases and symptoms without considering the duration it takes to develop. Most commonly reported symptoms and comorbid conditions are found enriched in our analysis at a cut off specific disease occurrence score of 4. Interestingly, some diseases/ symptoms reported in isolated COVID-19 cases are also found beyond the cut off 4 (**Fig-4**).

**Fig-4:**
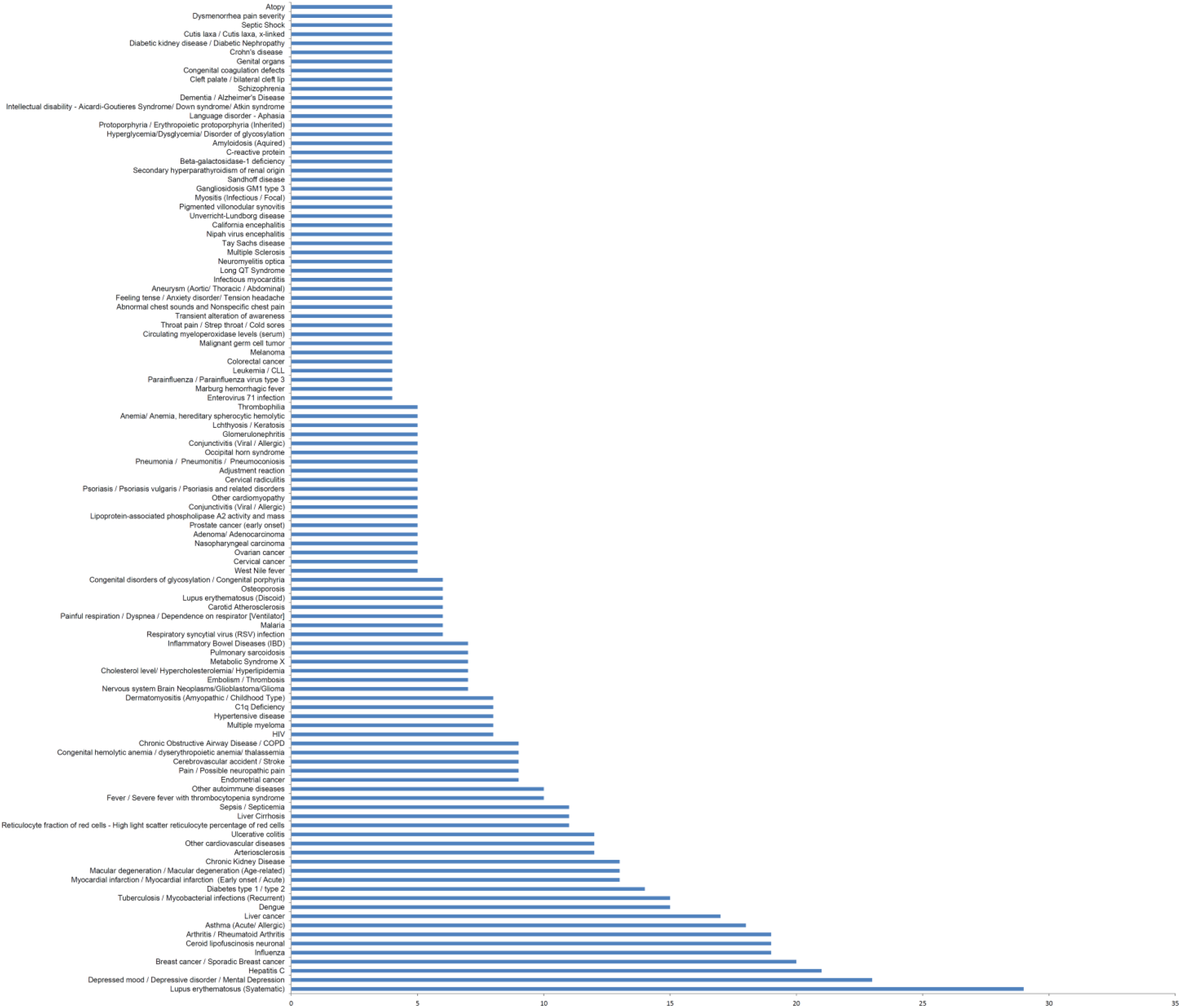
Top specific diseases without considering their duration to develop the clinical symptoms.

According to rank (with a cut off score 4), the enriched known or reported comorbid conditions or long-term complications are lupus erythematosus (Rank-1) [100], depressive disorder / mental depression (Rank-2) [30, 101], hepatitis C (Rank-3) [102], breast cancer (Rank-4) [75], arthritis (Rank-5) [103], asthma (Rank-6) [10, 77], liver cancer (Rank-6) [78], tuberculosis (Rank-7) [104, 105], diabetes (Rank-8) [82], myocardial infarction (Rank-9) [35, 106], macular degeneration (Rank-9) [80], chronic kidney disease (Rank-9) [56, 57], arteriosclerosis (Rank-10) [107], ulcerative colitis (Rank-10) [108], liver cirrhosis (Rank-11) [12, 109], sepsis / septicaemia /septic shock (Rank-11) [60, 81], other autoimmune diseases (Rank-12) [49, 50], cerebrovascular accident / stroke (Rank-13) [110], congenital anemia (Rank-13) [111], COPD (Rank-13) [86], multiple myeloma (Rank-14) [112], hypertensive disease (Rank-14) [9, 113], dyslipidemia (Rank-15) [93], metabolic syndrome X (Rank-15) [51, 114], osteoporosis (Rank-16) [91], prostate cancer (Rank-17) [115], psoriasis (Rank-17) [116], leukemia (Rank-18) [117, 118], long QT syndrome (Rank-18) [119], dementia (Rank-18) [120], schizophrenia (Rank-18) [121], intellectual disability (Rank-18) [122, 123], diabetic kidney disease/ diabetic nephropathy (Rank-18) [124]etc.

In our analysis other clinical conditions such as ceroid lipofuscinosis neuronal (Rank-5), endometrial cancer (Rank-13), C1q deficiency (Rank-14), brain tumor (Rank-15), pulmonary sarcoidosis (Rank-15), congenital disorders of glycosylation (Rank-16), cervical and ovarian cancers (Rank-17), nasopharyngeal carcinoma (Rank-17), glomerulonephritis (Rank-17), thrombophilia (Rank-17), and melanoma (Rank-18) are also enriched. However, we could not get any report associating these conditions with COVID-19 (**Fig-4**). As in our analysis most of the diseases are found correlated with COVID-19, we presume that these conditions could also be either comorbid or long-term consequences of COVID-19.

Further, in this analysis, we also found some diseases that are not so far reported to be associated with the increased risk or severity of COVID-19 such as HIV (Rank-14) [125], IBD (Rank-15) [95], and neuromyelitis optica (Rank-18) [126]. Interestingly, we also found atopy at Rank-18 which is found to have a protective role from severe complications of COVID-19 [127, 128].

In this analysis, we also found several common symptoms (mild, severe, and long-term complications) of COVID-19 are enriched at a cut off value 4. These include influenza (Rank-5) [129], dengue (Rank-7) [130], fever / severe fever with thrombocytopenia syndrome (Rank-12) [5], pain (Rank-13) [18], dermatomyositis (Rank-14) [131], embolism / thrombosis (Rank-15) [132], dyspnea / dependence on respirator (ventilator) (Rank-16) [5], conjunctivitis (Rank-17) [99], pneumonia / pneumonitis (Rank-17) [86], lchthyosis / keratosis (Rank-17) [133], throat pain / strep throat / cold sores (Rank-18) [134], fatigue/ transient alteration of awareness (Rank-18) [5], abnormal chest sounds and nonspecific chest pain (Rank-18) [134], anxiety disorder (Rank-18) [135], infectious myocarditis (Rank-18) [136, 137], aneurysm (Rank-18) [138], and myositis (Rank-18) [139], (**Fig-4**). However, we could not find any report of COVID-19 association with cervical radiculitis (Rank-17), adjustment reaction (Rank-17), and dysmenorrhea pain (Rank-18).

Similar to the previous analysis, in this ranking we also found many diseases/symptoms reportedly associated with COVID-19 below the cut off 4. For example lactate dehydrogenase level (Rank 21) and D-dimer level (Rank 21) [16] etc. For the full list of the specific diseases/symptoms please see the (**Fig-4**).

### COVID-19 associated specific diseases, symptoms, and complications based on their time to develop

For better understanding of the COVID-19 associated specific clinical conditions based on our multi-omics profiles, we classified the 403 specific disease or symptoms based on the time or duration it takes to develop and if genetics is also involved. We grouped the diseases under six categories: short, short-mid, mid - long, short - mid - long, short – mid - long (genetic), mid - long (genetic), and congenital/ genetic.

#### Short-term diseases or signs and symptoms

Under this group, we classified the diseases/ signs/ symptoms under two groups: infectious and non-infectious or non-communicable diseases (NCDs). We found 35 infectious diseases are enriched and the top five diseases at a cut off count of occurrence of 4 are influenza (Rank-1), dengue (Rank-2), respiratory syncytial virus (RSV) infection (Rank-3), malaria (Rank-3), West Nile fever (Rank-4), marburg hemorrhagic fever (Rank-5) etc. (**Fig-5A**). For the non-infectious disease/ NCDs or signs and symptoms group, we found 54 different short-term conditions. Among these, high light scatter reticulocyte percentage of red cells (Rank-1), sepsis or septicaemia (Rank-1), fever / severe fever with thrombocytopenia syndrome (Rank-2), pain (Rank-3), painful respiration or dyspnea or dependence on respirator (ventilator) (Rank-4), lipoprotein-associated phospholipase A2 activity and mass (Rank-5), conjunctivitis (Rank-5), pneumonia (Rank-5), thrombophilia (Rank-5), serum myeloperoxidase level (Rank-6), throat pain or strep throat (Rank-6), fatigue/ transient alteration of awareness (Rank-6), abnormal chest sounds and pain (Rank-6), anxiety and headache (Rank-6), viral encephalitis (Rank-6), C-reactive protein level (Rank-6), and septic shock (Rank-6) are the top conditions with the cut off occurrence score of 4. The details of these diseases or symptoms are presented in **Fig-5A**.

**Fig-5:**
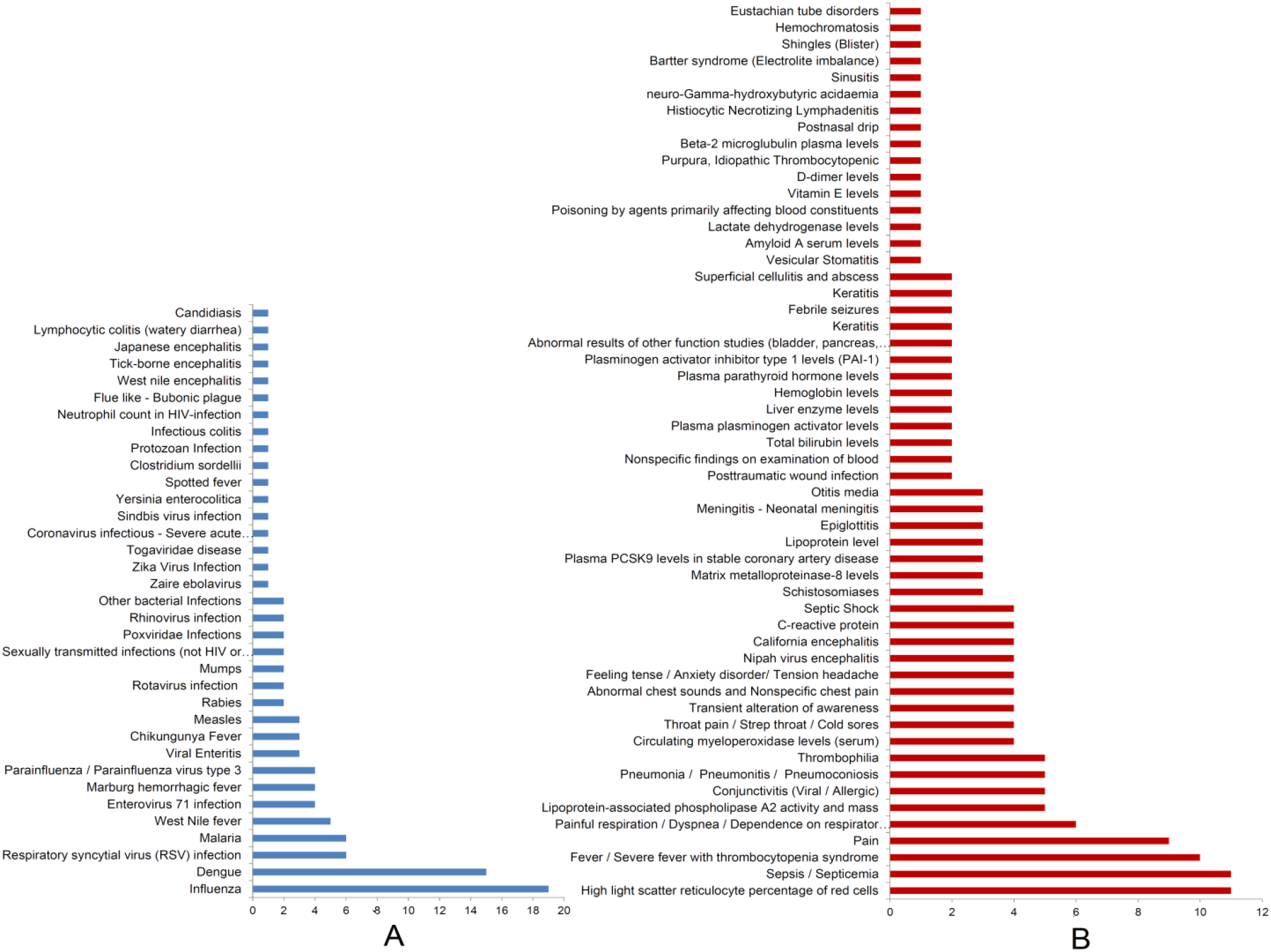
A) Short-term infectious diseases, B) Short-term NCDs/ signs/ symptoms

According to WHO, most common symptoms in COVID-19 are fever, dry cough and, tiredness. The less common symptoms include ache and pain, sore throat, diarrhoea, conjunctivitis, headache, loss of taste or smell, skin rash, and discolouration of toes or fingers. The serious symptoms that require medical interventions are breathing difficulty or shortness of breath, chest pain or pressure, and loss of speech or movement [140]. COVID-19 and influenza shares many common symptoms however, there are many differences too [141]. In our short-term infectious disease analysis, we observed that, influenza is the top candidate followed by dengue, respiratory syncytial virus (RSV) infection, and malaria (**Fig-5A**). Literature also suggests that COVID-19 shares symptoms of dengue [130] and differential diagnosis includes all kinds of respiratory viral infections including respiratory syncytial virus (RSV) [142]. Similarly, malaria also shares many important symptoms of COVID-19 [143]. Therefore, the viral infections enriched in our analysis are highly accurate and all the viral diseases flagged in our analysis need to be considered with differential diagnosis for COVID-19.

While we considered the short-term specific non-infectious diseases/NCDs/ signs / symptoms, we found that most of the COVID-19 symptoms as described by WHO [140] such as fever / severe fever with thrombocytopenia syndrome (Rank-2) [144], pain (Rank-3) [145], painful respiration / dyspnea / dependence on respirator [Ventilator] (Rank-4) [5], conjunctivitis (Rank-5) [99], pneumonia (Rank-5) [86], headache (Rank-6) [145, 146], transient alteration of awareness (tiredness/ fatigue) (Rank-6) [5], strep throat/ sore throat (Rank-6) [134], abnormal chest sounds and nonspecific chest pain (Rank-6) [134] are found in our top ranked symptoms (**Fig-5A**). However, dry cough, loss of taste or smell, and loss of speech or movement are not enriched as short-term symptoms. Interestingly, high light scatter reticulocyte percentage of red cells is found as the top candidate (Rank-1) sign in our analysis. Recently, high reticulocyte is reported to be significantly higher in critical patients and may be a prognostic predictor for poor outcomes [147]. Similarly, sepsis or septicemia is also enriched at Rank-1 in our analysis and sepsis is found to worsen the condition of critically ill COVID-19 patients [60].

The cardiovascular diseases biomarker lipoprotein-associated phospholipase A2 (PLA2G7) is highly upregulated in severe COVID-19 pneumonia [148]. In our result, we found the lipoprotein-associated phospholipase A2 activity is ranked at 5. Thrombophilia, which also came at Rank-5, is highly prevalent in COVID-19 patients, however, is not associated with disease severity [149]. Myeloperoxidase could be a potential biomarker for detection of SARS-CoV-2 infection [150]. The serum myeloperoxidase level is found enriched at Rank-6 along with viral encephalitis, C-reactive protein, and septic shock (**Fig-5A**). Encephalitis is frequent in older patients having age group >50 and mostly develops while under treatment in ICU [83], level of C-reactive protein is positively associated with lung lesions and disease severity in COVID-19 [151], and 70% of COVID-19 non-survivors develop septic shock [152]. Schistosomiases, matrix metalloproteinase level, plasma PCSK9 levels, lipoprotein level, epiglottitis, meningitis, and otitis media are found at Rank-7 (with the cut off occurrence score of 3) in our analysis. An early up-regulation of circulating MMP-9 is associated with respiratory failure in COVID-19 [153], and lipoprotein level is associated with severity of COVID-19 [93], while in September 2020 acute epiglottitis [154] and meningitis as initial symptoms of COVID-19 [155] are found in two separate COVID-19 patients. Similarly, otitis media is also observed in isolated cases [156]. Several conditions came at Rank-8 with a cut off occurrence score of 2. Among them, higher bilirubin level is observed in severe COVID-19 cases [157], plasminogen activator inhibitor type 1 (PAI-1) is upregulated and extremely high levels of plasminogen activator (tPA) enhance spontaneous fibrinolysis and mortality [158], and lower haemoglobin level is observed in severe COVID-19 cases [159]. Deregulation of parathyroid hormone level [160, 161], elevated liver enzyme levels [162], keratitis [163], febrile seizures [164, 165], cellulitis and sinusitis [166] are reported in isolated COVID-19 cases. Higher level of blood lactate dehydrogenase and D-dimer (Rank-9, score-1) is associated with disease severity in COVID-19 [16] (**Fig-5A**).

Therefore, ∼90% of our predicted short-term diseases/symptoms/conditions associated with COVID-19 are correct at an occurrence cut off of 4 and several conditions associated with COVID-19 are also found enriched beyond the occurrence cut off of 4 indicating the high accuracy of the method we have developed. Further, majority of these identified diseases/symptoms/conditions are associated with COVID-19 severity and disease outcomes. Therefore, we predict that the signs and symptoms that we have identified, but not discussed or so far not reported in the literature may also be associated with COVID-19.

#### Short - Mid and Short -Mid -Long term diseases or conditions

**Under short to mid conditions**, the top three conditions are hyperglycemia or dysglycemia (Rank-1), abnormal erythrocyte morphology (Rank-1), and chronic ulcer (Rank-1) (**Fig-6A**). COVID-19 is reported to aggravate dysglycemia and in diabetic patients, it leads to hyperglycemia and poor patient outcomes [167]. COVID-19 patients with sepsis shows adverse effects on red blood cell morphology [168], and re-bleeding from peptic ulcer is common in COVID-19 patients [169].

**Fig-6:**
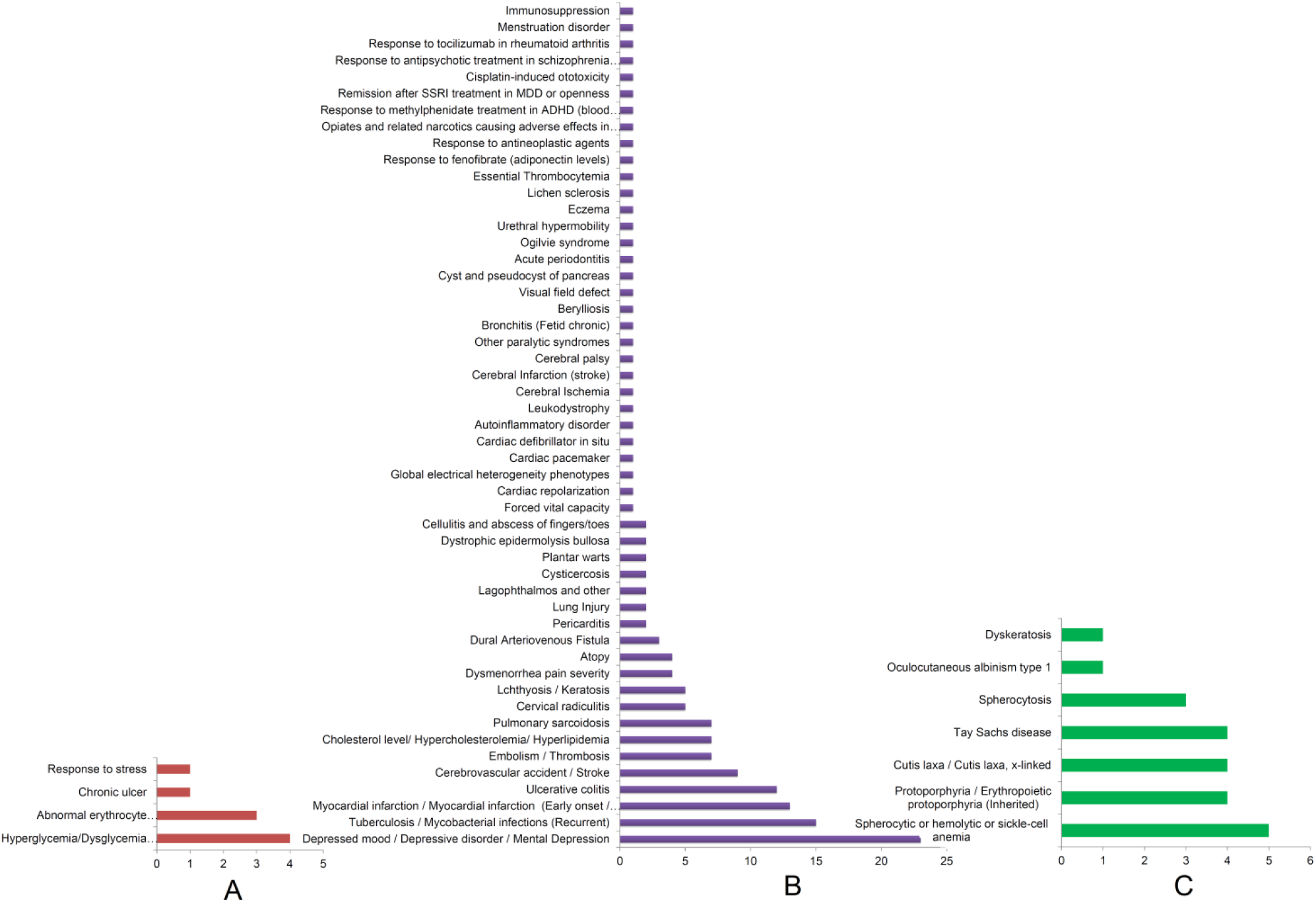
**A)** Short to mid specific NCDs / conditions. **B)** Short to mid to long specific NCDs / conditions. **C)** Short to mid to long (genetic) specific NCDs / conditions.

**Under Short to Mid to Long conditions**, the top five conditions found to be enriched are depressive disorders or depressed mood or mental depression at first rank (Rank-1) followed by recurrent mycobacterial infections (Rank-2), myocardial infarction (Rank-3), ulcerative colitis (Rank-4), and cerebrovascular accident or stroke (Rank-5). Embolism or thrombosis, cholesterol level, and pulmonary sarcoidosis came at Rank-6. Cervical radiculitis and lchthyosis or keratosis are found to enrich at Rank −7. Dysmenorrhea pain and atopy are ranked at Rank-8 with an occurrence cut off of 4 followed by dural arteriovenous fistula (Rank-9) with an occurrence cut off of 3. Pericarditis, lung injury, lagophthalmos, cysticercosis, plantar warts, dystrophic epidermolysis bullosa, and cellulitis and abscess of fingers/toes are enriched at Rank-10 with a cut off occurrence score of 2 (**Fig-6B**).

Current evidence suggests that COVID-19 shows various mental health problems including depression and mood and anxiety disorders [30, 101]. Nearly 28% COVID-19 patients are found to develop depressive mood disorders during their treatment [170] and acute COVID-19 patients may develop delirium in a significant proportion [171]. A recent report suggests that, a considerable percentage of COVID-19 patients develop mental disorders post recovery from the disease (14-90 days) and the patients having previous history of psychiatric disorders have higher incidence of COVID-19 [34]. Some reports suggest a higher mortality rate (11.6% to 14.3%) in patients with tuberculosis and COVID-19 co-infection [104, 105]. Myocardial infarction that came at Rank-3 is reported to be one of the most frequent cardiovascular syndromes in COVID-19 [35, 106] and in a meta-analysis ulcerative colitis (Rank-4) is found in 41.6% COVID-19 patients [108] and represents more severe symptoms than the patients with Crohn’s disease [172]. In a single centre study, although stroke (Rank-5) is observed in 4.6% [173] and in a meta-analysis 1.1% COVID-19 cases; COVID-19 patients having stroke show severe symptoms, poorer prognosis, and 46.7% mortality rate [110]. Pulmonary embolism (Rank-6) and deep vein thrombosis are reported in 20%-40% and 0% to 69% cases in COVID-19, respectively [132]. A sharp decrease in total cholesterol level (Rank-6) and low-density lipoprotein is observed in COVID-19 patients [92] and hypolipidemia correlates with COVID-19 severity [93]. Skin disease Keratosis that came at Rank-7 with an occurrence cut off of 5 is observed as a symptom in COVID-19 [133] and a prehistory of actinic keratosis exhibit 10.8% COVID-19 cases [116]. Acute pericarditis (Rank-10) is reported as a primary symptom in COVID-19 in some cases [174] and lung injury (Rank-10) during COVID-19 and subsequent pulmonary fibrosis in COVID-19 survivors are a frequent finding [175]. Interestingly, atopy that came at Rank 8 with cumulative score of 4 is reported to show positive effect in COVID-19. SARS-CoV-2 infected patients having prehistory of atopy require less hospitalization and showed decreased mortality [127] (**Fig-6B**). Similarly, menstruation has some protective effect from COVID-19 [176]. We could not find any literature showing correlation of COVID-19 with pulmonary sarcoidosis (Rank-6), cervical radiculitis (Rank-7), and dysmenorrhea pain severity (Rank-8). As in this analysis, we also found >90% precision at a cut off count of occurrence of 4, our other identified specific disease or symptoms could also be correlated with COVID-19 either in the form of a disease or comorbidity or long-term effect of COVID-19.

Since we have used OMIM Expanded and Rare Diseases databases in our Enricher-based [42] prediction, we got several **genetic diseases** as top candidates associated with COVID-19. These diseases are as follows: hereditary spherocytic/ haemolytic/ sickle cell anemia is the top disease (Rank-1) followed by erythropoietic protoporphyria (Rank-2), Cutis laxa, x-linked (Rank-2), Tay Sachs disease (Rank-2), spherocytosis (Rank-3), and dyskeratosis (Rank-4) (**Fig-6C**). COVID-19 severity is associated with anemia [90] and patients with sickle cell anemia are at higher risk of developing severe COVID-19 [111]. However, we don’t see any report on our other identified genetic diseases and their correlation with COVID-19. Nevertheless, based on the high confidence of our analysis, we presume that the identified genetic conditions need careful observation on their genetic disease - COVID-19 interactions and outcomes in the long term.

#### Mid – long term diseases or conditions

While we classified the mid – long term diseases or conditions into infectious, non-infectious, and genetics disease, four infectious diseases are enriched. These diseases are hepatitis C (Rank 1) followed by HIV (Rank 2), hepatitis B (Rank 3), and melioidosis (Rank 4) (**Fig-7A**). Hepatitis is recently found to be a comorbid condition in COVID-19. In a systematic review it is reported that 14.1% and 21.4% of COVID-19 patients with hepatitis B and hepatitis C require ICU admission and hepatitis C confers 13% increased risk of fatality in COVID-19 patients [102]. However, people living with HIV (PLHIV) do not show poorer outcomes from COVID-19 [125]. We did not get any report on correlation between melioidosis and COVID-19.

**Fig-7:**
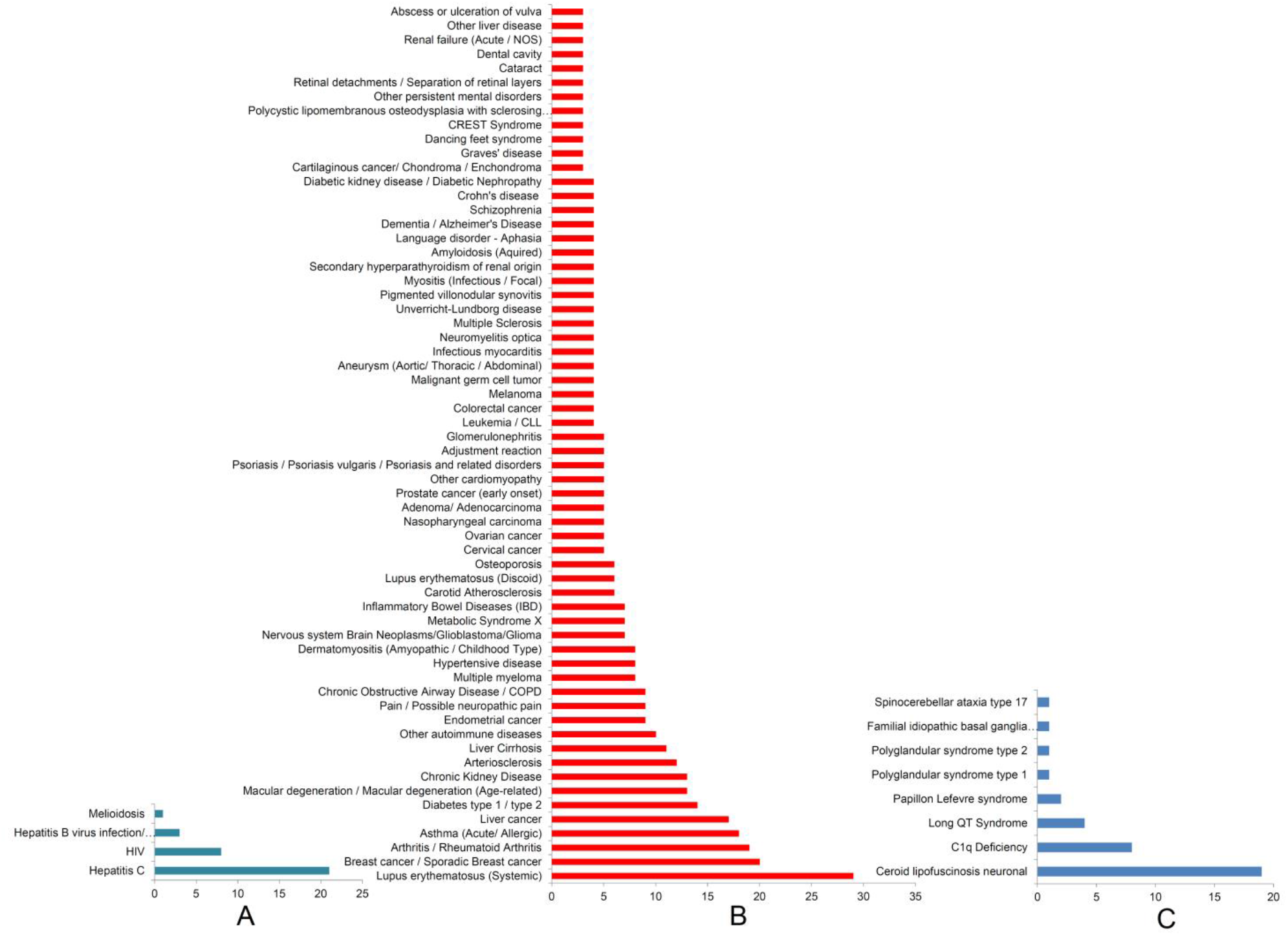
**A)** Mid to long specific infectious diseases. **B)** Mid to long specific NCDs / conditions. **C)** Mid to long (genetic) specific NCDs / conditions.

The top 17 non-infectious diseases or NCD conditions enriched at a cut off occurrence score of 4 as per their rank are systematic lupus erythematosus (Rank-1), breast cancer (Rank-2), arthritis or rheumatoid arthritis (Rank-3), asthma (Rank-4), liver cancer (Rank-5), and diabetes (Rank-6). At Rank-7 macular degeneration, and chronic kidney disease are enriched with a cut off score of 13. Arteriosclerosis, liver cirrhosis, and other autoimmune diseases came at Rank-8, −9, and −10, respectively. Endometrial cancer, pain/ neuropathic pain, and COPD are enriched at Rank-11. At Rank-12, three diseases are enriched with a cut off score of 8. These diseases are multiple myeloma, hypertension/ hypertensive disease, and dermatomyositis. Glioma, metabolic syndrome X, and inflammatory bowel disease are Ranked at 13. At Rank-14 carotid atherosclerosis, osteoporosis, and lupus erythematosus (discoid) are found. Nine diseases are ranked at 15. They are cervical cancer, ovarian cancer, nasopharyngeal carcinoma, prostate cancer, other cardiomyopathy, psoriasis, adjustment reaction, and glomerulonephritis. At Rank-16, nineteen diseases are enriched with a cut off score of 4. These diseases are leukemia, colorectal cancer, melanoma, malignant germ cell tumor, aneurysm, infectious myocarditis, neuromyelitis optica, multiple sclerosis, Unverricht-Lundborg disease, pigmented villonodular synovitis, myositis, secondary hyperparathyroidism of renal origin, acquired amyloidosis, aphasia, dementia or Alzheimer’s disease, schizophrenia, Crohn’s disease, and diabetic kidney disease / diabetic nephropathy. Several other conditions are also enriched and the full list is given in **Fig-7B**.

Literature suggests that lupus (Rank-1) patients are more vulnerable to COVID-19 [100] and breast cancer (Rank-2) patients infected with COVID-19 show 13% mortality rate [75]. Arthritis that came at Rank-3 is reported to increase risk and severity of COVID-19 [103]. Although asthma came at Rank-4 and a comorbid condition, it may not increase the mortality rate [10, 77]. Hepatic injury is observed in 10-40% of patients with COVID-19 and therefore, in liver cancer (Rank-5) patients, COVID-19 aggravates the disease severity [78]. Diabetes that came at Rank-6 is having a bidirectional association with COVID-19. Diabetes increases the risk of severe COVID-19 or COVID-q9 associated mortality and patients with COVID-19 show early onset diabetes and severe metabolic complications associated with the disease [33, 82, 113]. Patients having a history of macular degeneration and or chronic kidney disease (Rank-7) are at increased risk of COVID-19 associated morbidity and mortality [80, 177, 178]. Reports suggest that arteriosclerosis or coronary artery calcification at Rank-8 is associated with worse prognosis [107] and liver cirrhosis (Rank-9) is associated with increased deaths in COVID-19 [12, 109]. Similarly, other autoimmune disease (Rank-10) is also associated with increased risk of COVID-19 [50]. Neuropathic pain and COPD at Rank-11 are also found to associate with COVID-19. Reports suggest that nearly 2.3% of COVID-19 patients display neuropathic pain [179, 180] and COPD patients are at high risk of developing severe pneumonia and poor outcomes if infected with SARS-CoV-2 [86].

Multiple myeloma and hypertension enriched at Rank-12 are also comorbid conditions. Multiple myeloma is associated with severe COVID-19 [112] and hypertension is found in 46% cases and is associated with severity and mortality of COVID 19 [9, 48, 113]. Similarly, other metabolic syndromes (Rank-13) are also associated with severity and death of COVID 19 [51, 114]. Individuals suffering from osteoporosis (Rank-14) [91], cardiomyopathy (Rank-15) [15], and psoriasis (Rank-15) are at high risk of developing severe COVID-19 [91].

With a cut off occurrence score of 4 or (Rank-16), we found several conditions that are associated with COVID-19. Subarachnoid hemorrhage (SAH) is observed in COVID-9 patients and instability of aortic aneurysm is associated with SAH [138]. Myocarditis is one of the direct consequences (7% - 23%) of SARS-CoV-2 infection and is associated with higher degree of morbidity and mortality from COVID-19 [136, 137]. Neurological changes caused by COVID-19 show similarities with multiple sclerosis [181] and myositis is a manifestation of SARS-CoV-2 infection [139]. Amyloid microclots are found in the native plasma of COVID-19 [182] and in diabetic patients, serum amyloid A (SAA) level may be associated with the severity of COVID-19 [183]. Aphasia is found in COVID-19 patients who have developed encephalopathy or ischemic stroke [184, 185]. Recent literature suggests that people suffering from dementia or Alzheimer’s disease are at high risk of being infected and developing severe symptoms of COVID-19 [186, 187]. Similar to dementia, schizophrenia patients are also at higher risk of SARS-CoV-2 infection and poorer outcomes [121]. Although, patients with Crohn’s disease exhibits less severe symptoms as compared to ulcerative colitis [108, 172], patients with diabetic nephropathy shows >2 fold COVID-19 associated pneumonia and higher mortality as compared to other chronic kidney disease [124]. As mentioned earlier, two conditions IBD (Rank-13) and neuromyelitis optica (Rank-16) may not be associated with COVID-19 associated risk or severity [95, 126]. On the other hand, literature suggests that COVID-19 may trigger the development of dermatomyositis (Rank-12) [131] and Glioma (Rank-13) [188]. Similar to other analysis, we also found several COVID-19 associated conditions below the occurrence cut off value of 4. Interestingly, in this group at Rank-19, we found specific psychiatric disorders such as suicide tendency addiction. Recent report suggests that substance use disorders (SUD) or drug addicted people are at higher risk of being infected and develop severe COVID-19 [189]. Similarly, COVID-19 positive patient without a pre-history of any psychiatric condition is reported to show confusion, psychotic symptoms, and suicide attempt [190]. Therefore, in this analysis also we achieved ∼90% accuracy in identifying comorbid or long-term consequences of COVID-19.

Importantly, we observed a lot of cancers enriched in our analysis such as breast cancer (Rank-2), liver cancer (Rank-5), endometrial cancer (Rank-11), cervical cancer, ovarian cancer, nasopharyngeal carcinoma, prostate cancer (Rank-15), leukemia, colorectal cancer, melanoma, malignant germ cell tumor (Rank-16), chondroma (Rank-17), sarcoma, lymphoma, larynx cancer (Rank-18), esophagal cancer, pharynx cancer, squamous cell carcinoma of mouth (Rank-19) among others (**Fig-7B**). Considering our analysis that is showing high confidence in identifying the COVID-19 related conditions and complications, coupled with the facts that (i) some of these cancers such as breast cancer [75], liver cancer [78], gynaecologic cancers [76], hematologic [79], colon cancer [87], prostate cancer [115]are already reported as comorbid conditions, (ii) glioma may be developed due to COVID-19 [188], and the fact of viral carcinogenesis process[191, 192]; we hypothesize that the cancers we have identified in our analysis could be the long-term consequence of COVID-19.

The genetic diseases that are identified for mid to long term based analysis are ceroid lipofuscinosis neuronal (Rank-1), C1q deficiency (Rank-2), long QT syndrome (Rank-3) Papillon Lefevre syndrome (Rank-3), Polyglandular syndrome (Rank-5), familial idiopathic basal ganglia calcification (Rank-5), and spinocerebellar ataxia (Rank-5) (**Fig-7C**). We could not find any literature correlating COVID-19 with these genetic conditions. Since our analysis is giving high accuracy in identifying COVID-19 associated conditions, a careful and long observation is required to understand if there is any association of SARS-CoV-2 infection with these genetic conditions.

### Congenital and genetic diseases

Interestingly in our analysis, we found 57 congenital or genetic diseases that could be related to COVID-19. The top three diseases are congenital hemolytic anemia (Rank-1), congenital disorders of glycosylation (Rank-2), and occipital horn syndrome (Rank-3). At Rank-4 with an occurrence cut off score of 4, we found seven diseases and they are gangliosidosis GM1 type 3, Sandhoff disease, beta-galactosidase-1 deficiency, syndrome associated intellectual disability, cleft palate, congenital coagulation defects, and genital organ defects. With an occurrence cut off score of 3, ceroid storage disease and aspartylglucosaminuria are enriched at Rank-5. Adrenoleukodystrophy, paget disease of bone, asperger syndrome, autism spectrum disorder, microcephaly, arthrogryposis, congenital limbs defect, cystic fibrosis, rhizomelic chondrodysplasia punctate, fetal hemoglobin quantitative trait locus, and familial erythrocytosis are found at Rank-6 with an occurrence cut off score of 2. The complete list of this group of diseases is given in **Fig-8**).

**Fig-8:**
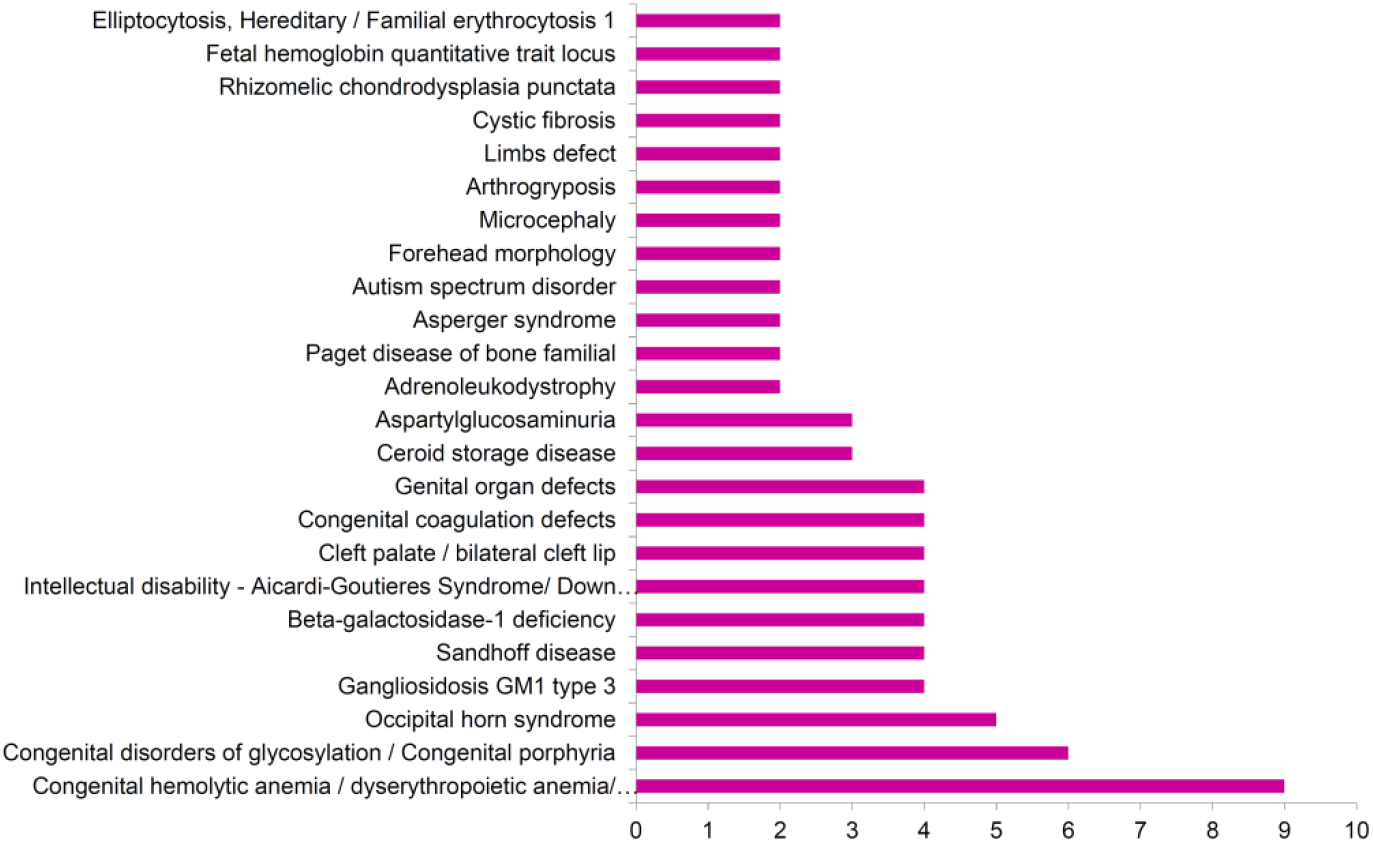
Congenital and genetic diseases associated with COVID-19.

Among these identified diseases, some congenital diseases have been reported to show positive or negative correlation with COVID-19. However, majority of the congenital diseases identified here have no information so far on their association with COVID-19. Patients with hemolytic anemia (Rank-1), specifically sickle cell anemia are at higher risk of developing severe COVID-19 [111]. Down syndrome (Rank-4) that represents intellectual disability exhibits 4-fold increased risk of severity and 10-fold increased risk of mortality from COVID-19 [193] and autism spectrum disorder (Rank-6) may also be a risk factor [88, 89]. On the other hand, congenital coagulation defects (Rank-4) or congenital bleeding disorders are associated with more favourable outcomes of COVID-19 [194] and fetal hemoglobin (Rank-6) may play a protective role against COVID-19 as the neonates, infants, and children show low prevalence of SARS-CoV-2 infection [195, 196]. Cystic fibrosis (Rank-6) patients may be at risk but are probably not vulnerable to COVID-19 due to their current treatment of cystic fibrosis [197, 198] and according to Paget’s Association, Paget’s disease (Rank-6) is not a risk factor for COVID-19 [199]. As our analysis is providing some important congenital disease and COVID-19 associations, the effect of COVID-19 on our other identified congenital conditions require careful and long-term investigation to find if there is any correlation.

### Long-term consequences of COVID-19

WHO predicted that COVID-19 could cause long-term health complications such as cardiac muscle damage and heart failure, damage of lung tissue and lung failure, anosmia, pulmonary embolism, heart attack, stroke, cognitive impairment, anxiety, depression, post-traumatic stress disorder, sleep disorder, pain in muscles and joint, fatigue [31]. A recent report suggest that nearly 70% young and low-risk COVID-19 survivors develop one or more organs impairment within four months after initial symptoms of SARS-CoV-2 infection. Impairment of heart is observed in 32% cases, lungs (33%), kidneys (12%), liver (10%), pancreas (17%), and spleen (6%). Further, single organ damage is observed in 66% cases while multi-organ injury is found in 25% subjects [32]. It is also reported recently that one in five COVID-19 survivors develops mental illness for the first time such as anxiety, depression, and insomnia within 90 days from the first COVID-19 symptoms shown and COVID-19 patients having pre-existing psychiatric disorder are at risk of COVID-19 severity [34]. The individuals recovering from COVID-19 are also developing substance use disorder [22], confusion, psychotic symptoms, and suicide tendency [30, 190]. Apart from these, COVID-19 is found to trigger arthritis [200], dermatomyositis [131], glioma [188], and early onset of diabetes and severe metabolic complications [33]. Considering the facts that (i) all these NCDs and mental health conditions are found enriched at top ranks in our analysis along with several conditions, (ii) high confidence of our analysis (∼90%), (iii) most of our identified conditions are associated with severity of COVID-19, (iv) the bidirectional relationship between diabetes and COVID-19 [33], psychiatric disorder and COVID-19 [34], COVID-19 and cardiovascular system diseases [35], (v) presence of SARS-CoV-2 viral RNA in patients body for long time after recovery [20], and (vi) our “genetic remittance” assumption [43]; we hypothesize that in long-term, COVID survivors may develop various conditions we have identified in this analysis. However, as COVID-19 is a new disease, long-term observational studies are required on COVID-19 survivors to assess the long-term consequences of the SARS-CoV-2 infection.

## CONCLUSION

In this analysis, using multi-omics (proteome, transcriptome, interactome, and bibliome) data of SARS-CoV-2 infection and a unique bioinformatics strategy, we have identified mild and severe symptoms, associated comorbid conditions, short and long term complications of COVID-19. Based on available literature, our predictions are found to have ∼90% precision in identifying the COVID-19 associated mild and severe symptoms, comorbid conditions, and long-haul complications. This indicates that the method we have developed is highly accurate and can be applied to other diseases and pandemics too. In this analysis, we have identified the disease/conditions using the omics profiles upregulated due to SARS-CoV-2 infection. Since the viral RNA remains active for several months within the COVID-19 survivors, these upregulated omics profile remains switched on for a long time and hence the corresponding NCD pathways may lead to the development of the respective NCD as a long-term consequence of SARS-CoV-2 infection. The limited literature on long-term consequences of COVID-19 so far available also supports our hypothesis. However, long-term observational studies are required to further support our findings.

## Data Availability

The source of data is mentioned in the article

## Author contribution

DB: conceived and designed the experiment, data collection and analysis, result interpretation, and wrote the paper; ST and BSA preformed re-analysis; PG: data interpretation and edited the article, MEW, AGN, VA, NKG: provided technical inputs. All authors have read and approved the article.

## Funding

None

## Competing interest

Authors declare no competing interest.

## Notes

### Competing Interest Statement

The authors have declared no competing interest.

### Author Declarations

The article does not require any IRB/oversight body approval

